# Body-focused repetitive behaviours in adolescents: a common and under-recognised source of distress and unmet need

**DOI:** 10.64898/2026.05.18.26353292

**Authors:** Clare Mackay, Polly Waite, Laura Lee, Holly Haines, Matthew Toher, OxWell Study Team, Mina Fazel

## Abstract

**Background:** Body-focused repetitive behaviours (BFRBs), including hair pulling, skin picking and nail biting, are common but under-recognised behaviours that often emerge during adolescence. Their prevalence, associated distress, and relationship with mental health and social factors remain poorly characterised.

**Methods:** 5,437 adolescents aged 11–18 years reported engagement in BFRBs, associated distress, and functional interference as part of the 2025 OxWell Student Survey. Participants. Problematic BFRBs were defined as at least one BFRB with moderate or high distress. Associations with gender, neurodivergence, bullying, and internalising symptoms were examined using multivariable logistic regression.

**Results:** Overall, 58.5% of participants reported at least one BFRB. Nail biting was most common (43.6%), followed by skin picking (31.3%) and hair pulling (14.5%). Among those with BFRBs, 22.2% reported moderate-to-high distress, and 3.3% of the total sample reported the highest level of distress. Co-occurrence was common: more than half of those with BFRBs reported multiple behaviours. BFRBs were more common and more distressing in girls and trans/gender-diverse participants than in boys. Problematic BFRBs showed strong associations with internalising symptoms (6.3% in the normal range vs 34.2% in the clinical range) and bullying (9.6% with no bullying vs 27.1% with >weekly bullying). Internalising symptoms were the strongest predictor in multivariable models (OR 1.97 per 10-point increase), alongside independent contributions from gender and frequent bullying.

**Conclusions:** BFRBs are common in adolescents, frequently co-occur, and are strongly associated with emotional distress and social adversity. Recognition of distress, rather than behaviour alone, may be important for identifying unmet need and guiding intervention.

## Introduction

Body-Focused Repetitive Behaviours (BFRBs) include actions such as picking, pulling, or biting at one’s hair, skin, or nails. While these are common behaviours within normal grooming practices, for some individuals they become compulsive, resulting in unintentional physical damage such as bald patches; loss of eyelashes, eyebrows or beard hair; skin lesions; and infections [1]. Although severe physical complications are relatively uncommon, the psychological impact can be substantial [2], [3]. These complex, multifactorial behaviours often emerge in childhood or adolescence [4] and are thought to be maintained through self-regulatory loops [5]. Despite this, BFRBs remain under-recognised in clinical, educational, and social contexts, and there is a gap in our understanding about the relationship between prevalence and psychosocial impact, especially in young people.

The existing literature generally focuses on individual BFRBs in isolation, limiting our understanding of their co-occurrence, overall prevalence, gender differences, and how they relate to other aspects of wellbeing. While BFRBs are classified within obsessive–compulsive and related disorders in both ICD-11 and DSM-5, the DSM-5 provides more explicit diagnostic definitions such that hair-pulling and skin-picking are formally recognised as trichotillomania and excoriation disorder, respectively (American Psychiatric Association, 2013). In contrast, behaviours such as nail-biting (onychophagia) and other repetitive grooming behaviours (e.g. lip-picking, cheek-biting, nose-picking) are not classified as distinct disorders in either system, but are instead captured under an ‘other specified’ category despite their overlapping psychological and behavioural features (American Psychiatric Association, 2013). Across types, BFRBs involve recurrent contact with one’s own body surface, sometimes preceded by tension or sensory discomfort and usually followed by transient relief [6]. Studies in adult samples using diagnostic tools typically report estimates of around 1–5% for hair-pulling and skin-picking disorders, and clinical studies have a marked female predominance of approximately 4:1 [1] [7] [8]. In contrast, larger self-report surveys conducted in web- or college-based samples yield substantially higher rates of 10–25% for non-clinical hair-pulling or skin-picking and up to 40–50% for nail-biting, with much smaller sex differences [9], [10], [11], [12]. Such discrepancies indicate that BFRBs exist along a continuum from common grooming behaviours to clinically impairing conditions, and we do not have a clear picture of how many people experience these behaviours as distressing, impairing, or are in need of support. Without this knowledge, the true size and shape of unmet need remains obscured, and opportunities for early recognition and intervention may be missed.

This is particularly pertinent during adolescence, which is the most common age of onset for BFRBs [13] and is a critical developmental period characterised by rapid changes in emotional regulation, peer relationships, and identity formation [14], [15]. The small number of prevalence studies in young people report ranges comparable to those observed in adults: 2– 10% for hair-pulling [16] [17], 8–25% for skin-picking [16]; and 30–40% for nail-biting [12], [16]. Co-occurring mental-health difficulties, including anxiety disorders and depression, are common, alongside elevated rates of neurodivergent conditions such as ADHD and autism [17]. Social factors also appear to play an important role; experiences such as bullying, exclusion, and a low sense of belonging can heighten self-consciousness and shame, potentially reinforcing these behaviours [18], [19]. Despite this, few studies have explored these interconnections at scale, particularly among gender-diverse and trans adolescents, who remain under-represented in prevalence research in general [20], and for whom there are no studies relating to BFRBs.

The OxWell Student Survey provides a unique opportunity to address these gaps. OxWell is a large, school-based survey of mental health and wellbeing conducted biannually in England [21]. For the 2025 wave, we introduced questions about BFRBs, co-designed with individuals who have lived experience of these behaviours, into an optional extra module offered to a subset of students from age 11 -18 years [22]. We assessed engagement in hair, skin-, and nail-focused behaviours experienced as difficult to control, and their associated distress and interference across key domains of functioning, including home life, friendships, leisure, and classroom learning. By integrating these items alongside validated measures of anxiety, depression, neurodiversity, and social context, we were able to: (1) estimate the proportion of the sample who experience the most common BFRBs (hair-pulling, skin-picking and nail-biting), and examine their co-occurrence patterns; (2) quantify associated distress and functional interference across multiple life domains; (3) examine gender differences in the prevalence and distress associated with BFRBs; and (4) test the hypothesis that problematic BFRBs will be more common in those with self-reported neurodivergence, experience of bullying, and higher levels of depression/anxiety.

By situating BFRBs within the broader ecology of adolescent mental health, this study seeks to advance understanding of the prevalence, correlates, and associated distress and impairment with these common yet overlooked behaviours. In turn, this will inform more inclusive approaches to recognition, screening, and support.

## Methods

### Design and Setting

This study was a cross-sectional observational analysis of data from the 2025 OxWell Student Survey, which was administered to secondary-school and further education college students across England, and included 32,102 pupils [22]. Participation in OxWell is voluntary and uncompensated, and most students completed the survey online as part of a lesson during the school day. At the end of the main survey, a subset of students were asked if they could be happy to complete a further optional survey. Ethical approval for the 2025 wave was granted by the University of Oxford Research Ethics Committee (R62366/RE017). Data for the present analyses were collected between January – April 2025. Analyses were performed in R (version 4.5.2) using packages including dplyr, ggplot2, and MASS.

### Participants

All students aged 11–18 years who provided informed consent and completed the optional BFRB module were eligible for inclusion. Of the 16,861 students who answered the final question on the core survey, 6,910 (41%) continued to the ‘optional’ survey. A small number of participants (N=70) aged 19 years or older were excluded to align the analytic sample with a typical school-age population.

### Measures

#### Demographic variables

Age was derived from a categorical survey item and converted to a continuous numeric variable (years) for analyses. Gender response options were: male, female, trans and gender diverse, prefer not to say, and not sure. For analysis we report descriptive results for all response categories. For regression modelling, gender was recoded into three categories: boys/men, girls/women, and trans/gender diverse/unsure (henceforth referred to as TGD). Responses of ‘prefer not to say’ and other non-informative or missing responses were retained for descriptive tables and treated as missing in models when necessary.

#### Body-focused repetitive behaviours (BFRBs)

BFRBs were assessed using three questions developed by the authors with input from 26 people with self-declared lived experience of BFRBs who trialed and commented on the items. The items were:

1. **Do you sometimes pick, pull or bite at your hair, skin or nails in a way that feels difficult to control?** Response (tick all that apply): No; Hair pulling; Skin picking; Nail biting; Other
2. **Does this behaviour upset or distress you?** Response: Not at all; Only a little; Quite a lot; A great deal
3. **Does this behaviour interfere with your everyday life in the following areas?** (a) Home life; (b) Friendships; (c) Leisure activities; (d) Classroom learning. Response for each domain: Not at all; Only a little; Quite a lot; A great deal

Binary indicator variables were created for each specific BFRB (hair pulling, skin picking, nail biting, other). Participants could endorse multiple BFRBs; indicators were therefore treated as non-mutually-exclusive. Where participants endorsed any specific BFRB, that behaviour was considered present even if the respondent also selected “No” (i.e. as more than one option could be chosen, presence of a behaviour overrode a simultaneous “No” tick). A total BFRB count variable was computed as the sum of the four indicators, and a separate indicator denoted absence of any BFRB (count = 0).

Distress was coded on a 0–3 ordered numeric scale corresponding to “Not at all” (0) through to “A great deal” (3) and treated as an ordered categorical outcome in ordinal regression models. Analyses of distress distributions were restricted to participants reporting at least one BFRB and providing valid distress responses.

**Problematic BFRBs (primary binary outcome):** in the absence of diagnostic interviews, participants were classified as having problematic BFRBs if they endorsed at least one BFRB and reported associated distress of “Quite a lot” or “A great deal” (i.e., distress score ≥2).

**Severity variable:** a three-level ordinal severity outcome was derived for secondary analyses: none (no BFRB), BFRB with low/no distress, and BFRB with high distress (problematic as defined above). For descriptive figures we also derived commonly used groupings such as “nail biting only” (nail biting endorsed and no other BFRB endorsed) and “three+” (endorsement of three or more BFRBs).

Survey-specific non-response categories (e.g. “NoResponse(Item)”) were recoded as missing throughout.

##### Functional interference

Interference in four domains (home life, friendships, leisure activities, classroom learning) was captured using the same four-point scale as distress. Where multiple responses were recorded within a domain or participants provided composite answers, the maximum reported severity for that domain was used. Domain variables were treated as ordered categorical variables in descriptive and plotted summaries.

#### Neurodivergence and autistic traits

Neurodivergence was derived from self-reported diagnoses of autism, ADHD, and other neurodevelopmental conditions and was simplified into four categories for analysis: none, autism, ADHD, and both autism+ADHD (those who endorsed ‘other’ without also endorsing autism or ADHD were not included).

Autistic traits were measured using six items drawn from the Comprehensive Autistic Trait Inventory (CATI, [23]) representing social interaction, communication/social camouflage, repetitive behaviours, cognitive rigidity, and sensory sensitivity. Items were rated 1–5, with the social interaction item reverse-coded to align directionality. Item scores were summed to produce a total (range 6–30). Where a single CATI item was missing the total score was prorated based on the mean of available items; participants missing two or more CATI items were excluded from CATI analyses. The CATI total was standardised (z-score) for models that included it. This abbreviated 6-item version of the CATI - approved by the original measure’s creators - has been used in another large-scale UK survey (the Born in Bradford study). However, it demonstrated suboptimal internal consistency in the present sample (Cronbach’s α = 0.63) and as it has not yet been psychometrically validated, results should be interpreted with caution [24] [25]).

#### Bullying

Recent bullying was assessed with a categorical item taken from the Revised Olweus Bullying Questionnaire [26] about frequency in the past couple of months. For analysis responses were collapsed into three categories: None (no bullying), **<weekly** (responses “Once, twice or a few times” and “2 or 3 times a month”), and **>weekly** (responses “About once a week” and “Several times a week”). Non-response categories were coded as missing.

#### Mental health (internalising symptoms)

Internalising symptoms were indexed using the Revised Children’s Anxiety and Depression Scale – 25 item version (RCADS [27]). For descriptive figures we used the RCADS total imputed category (‘normal’, ‘borderline’ or ‘clinical’). The primary multivariable model reported in the manuscript includes the RCADS total imputed score entered as a continuous predictor. In post-hoc analyses depression and anxiety subscale scores were entered simultaneously to examine their independent associations, interpreted cautiously because of their high correlation.

### Statistical analysis

All analyses were performed in R (version 4.5.2). Primary data cleaning and aggregation used dplyr; figures used ggplot2; regression models used MASS (polr) and stats::glm.

#### Descriptive analyses

Point-frequency estimates were calculated for each BFRB type and for the composite indicators. Tabulations and stacked proportion plots were used to describe distress and interference within BFRB groupings and by gender.

To examine the extent to which the optional BFRB module sample differed from the full OxWell cohort, gender-standardised prevalence was obtained by weighting gender specific prevalence in the analytic sample to the gender distribution of the full cohort.

Co-occurrence of BFRBs was examined using the total number of endorsed behaviours (BFRB count). The proportion of participants reporting multiple BFRBs (≥2) was calculated, and conditional probabilities were used to compare the likelihood of multiple behaviours among participants endorsing specific BFRB types.

#### Distress (ordinal) analyses

Variation in distress by BFRB type was examined using proportional odds ordinal logistic regression (MASS::polr), with the ordered distress outcome (0–3). Non-mutually exclusive binary indicators for hair pulling, skin picking, nail biting, and other BFRBs were entered simultaneously, with gender included as an adjustment covariate. Predicted probabilities from these fitted models were computed across combinations of BFRB indicators to aid interpretation.

**Assumption:** proportional odds models assume that predictor effects are consistent across cumulative thresholds of the outcome; this assumption was acknowledged when interpreting results.

#### Primary model — problematic BFRBs

Problematic BFRBs (binary outcome) were analysed using multivariable logistic regression (binomial family with logit link). Predictors entered simultaneously were gender, neurodivergence, bullying frequency, RCADS total imputed score, and age. Boys/men, no neurodivergence, and no bullying served as reference categories. Adjusted odds ratios (ORs) and 95% confidence intervals (CIs) were obtained by exponentiating model coefficients and standard errors. Observations with missing data on any model variable were excluded from the fitted model.

Analyses did not explicitly account for clustering at the school level; given our focus on population-level associations and the large sample size we did not model clustering in primary analyses.

Adjusted odds ratios and 95% Wald confidence intervals were obtained by exponentiation of coefficients and their standard errors.

#### Secondary severity and post-hoc analyses

BFRB severity (three-level ordinal) was analysed using proportional odds regression as a secondary analysis. Post-hoc analyses included: (1) models entering depression and anxiety subscale scores simultaneously to assess independent effects; and (2) models that included both the CATI total and categorical neurodivergence to assess whether dimensional autistic traits attenuated associations with categorical neurodivergence.

#### Missing data

Analyses used available-case (complete-case) approaches per model: observations missing any variable required by a particular model were excluded from that model. Survey specific non-response codes (e.g. NoResponse(Item), NoResponse(Stopped_vB)) were treated as missing in all analyses.

### Statistical significance

Statistical significance was defined as p < .05. All tests are two sided.

## Results

A total of 5,437 OxWell 2025 participants aged 11-18 years completed the optional BFRB questions and are included in this analysis. Demographic, BFRB and covariate summary data are shown in Table 1. Age data were missing for 217 participants due to item non-response. Compared with the full OxWell 2025 sample (N=32,102), the BFRB subset included a greater proportion of girls/women (55% vs 46%) and trans/diverse/unsure participants (4.8% vs 3.5%), and a lower proportion of boys/men (38% vs 44.8%) (X-sq=491.7, df=3, p<0.01). Although there was a statistically significant difference in age between participants who did and did not complete the optional BFRB items (p < .001), the magnitude of this difference was negligible (M = 14.1 vs 13.8 years; Cohen’s d = 0.12).

**Table 1.** demographics and summary measures.

| variable | Boys/men | Girls/women | TGD | Total |
| --- | --- | --- | --- | --- |
| N | 2112 | 3132 | 351 | 5437 |
| Age in years, mean (SD) | 13.7 (1.8) | 13.9 (1.8) | 14.7 (2.1) | 13.8 (1.8) |
| Any BFRB N (%) | 928 (45.6%) | 1973 (64.6%) | 229 (84.2%) | 3183 (58.5%) |
| Hair pulling | 133 (6.5%) | 538 (17.6%) | 92 (33.8%) | 787 (14.5%) |
| Skin picking | 305 (15.0%) | 1191 (39.0%) | 173 (63.6%) | 1704 (31.3%) |
| Nail biting | 759 (37.3%) | 1424 (46.6%) | 156 (57.4%) | 2369 (43.6%) |
| Problematic BFRBs<br>(distress at least 'quite a lot') | 113 (5.6%) | 500 (16.4%) | 68 (25.0%) | 691 (12.7%) |
| RCADS total mean (SD) | 16.4 (12.9) | 26.2 (15.4) | 37.8 (15.8) | 23.1 (15.6) |
| Bullying N (%) |  |  |  |  |
| None | 1528 (77.6%) | 2292 (77.4%) | 140 (54.1%) | 3999 (76.2%) |
| <weekly | 308 (15.6%) | 516 (17.4%) | 78 (30.1%) | 916 (17.5%) |
| >weekly | 134 (6.8%) | 152 (5.1%) | 41 (15.8%) | 332 (6.3%) |
| Neurodivergence: |  |  |  |  |
| None | 1471 (73.4%) | 2327 (78.4%) | 112 (42.1%) | 3968 (74.7%) |
| Autism | 158 (7.9%) | 145 (4.9%) | 48 (18.0%) | 358 (6.7%) |
| ADHD | 219 (10.9%) | 331 (11.2%) | 30 (11.3%) | 585 (11.0%) |
| Both | 155 (7.7%) | 164 (5.5%) | 76 (28.6%) | 401 (7.5%) |
| CATI mean (SD) | 17.9 (4.5) | 19.5 (4.4) | 23.0 (4.0) | 19.1 (4.6) |

### Prevalence and co-occurrence patterns

Overall, 58.5% of participants reported engaging in at least one BFRB (Figure 1a). Nail biting was the most commonly reported behaviour (43.6%), followed by skin picking (31.3%) and hair pulling (14.5%), while 6.6% reported other BFRBs. Participants could endorse multiple behaviours; therefore, proportions for individual BFRBs are not mutually exclusive. Given the overrepresentation of girls/women and TGD participants in the BFRB sample, a gender-standardised estimate of the proportion of BFRBs in the full OxWell cohort was slightly lower at 56.9%.

**Figure 1.**
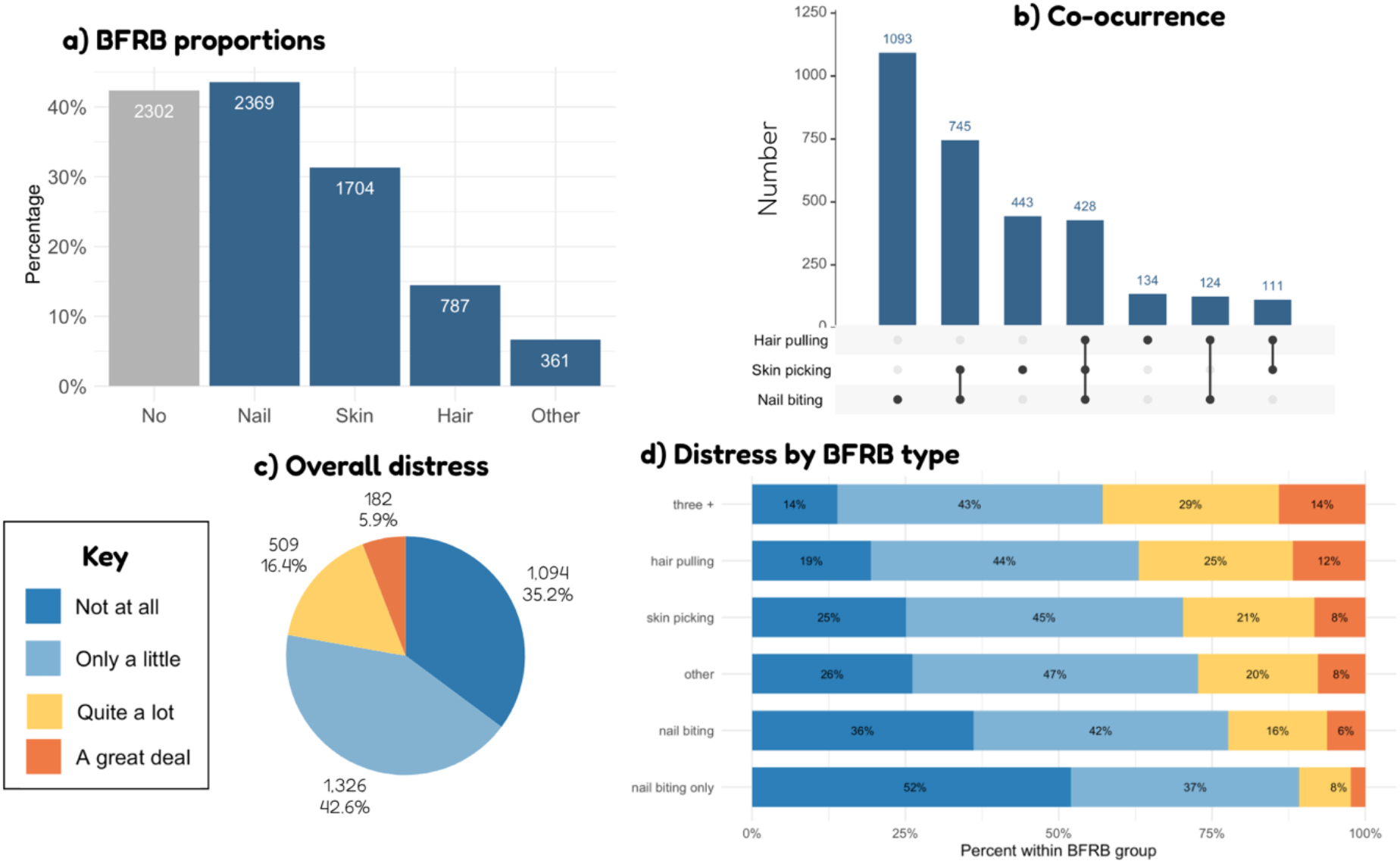
(a) Overall proportion of BFRBs in the sample of OxWell 2025 participants who undertook the BFRB questions. (b) UpSet plot of BFRB combinations, showing that while nail biting often occurred in isolation, hair pulling rarely did. (c) Overall distress levels show that while most people are not distressed by BFRBs, a significant minority are. (d) Distress by BFRB type, showing that greater distress is associated with hair-pulling and skin-picking than nail-biting, and greatest distress is associated with having multiple BFRBs.

BFRBs frequently co-occurred within individuals. While 31.8% of participants reported a single behaviour, 17.6% reported two behaviours, 7.7% reported three behaviours, and 1.5% reported all four behaviours. Among those reporting any BFRB, nearly half reported more than one behaviour, indicating that these behaviours commonly cluster rather than occurring in isolation. Co-occurrence also varied by BFRB type: among participants reporting hair pulling, 83.6% also reported at least one additional BFRB, compared with 75.6% of those reporting skin picking and 55.8% of those reporting nail biting (Figure 1b).

### Distress and functional interference

Among participants reporting at least one BFRB and providing valid distress data, most described low levels of associated distress, with 35.2% reporting no distress and 42.6% reporting only a little distress. However, a substantial minority reported higher levels of distress, with 16.4% reporting “quite a lot” and 5.9% reporting “a great deal”, indicating that 22.2% experienced moderate to high levels of distress (Figure 1c). Taken as a proportion of the whole sample, 691 participants (12.7%) met criteria for problematic BFRBs, defined as reporting at least one BFRB together with moderate to high levels of distress, and 182 participants (3.3%) reported the highest level of distress.

Distress varied by BFRB type (Figure 1d). Nail biting alone was associated with relatively low levels of distress, with the majority of participants reporting no or only minimal distress. In contrast, hair pulling and skin picking were associated with substantially higher levels of distress, with a greater proportion of participants reporting “quite a lot” or “a great deal” of distress. Participants reporting multiple BFRBs (three or more) showed the highest levels of distress overall.

In ordinal regression analyses adjusting for co-occurring BFRBs and gender, hair pulling and skin picking were both associated with substantially higher levels of distress (hair pulling: OR = 2.32, 95% CI 1.98–2.71; skin picking: OR = 2.22, 95% CI 1.92–2.56). Nail biting was associated with a more modest increase in distress (OR = 1.29, 95% CI 1.11–1.51), while other BFRBs showed intermediate effects (OR = 1.64, 95% CI 1.33–2.02).

BFRBs were associated with functional interference across multiple domains, including home life, friendships, leisure activities, and classroom learning (Figure 2). Interference was generally low among those reporting nail biting alone but increased markedly among those reporting skin picking, hair pulling and among those experiencing the highest levels of distress. Across all domains, participants reporting a great deal of distress also showed the greatest levels of functional impairment.

**Figure 2.**
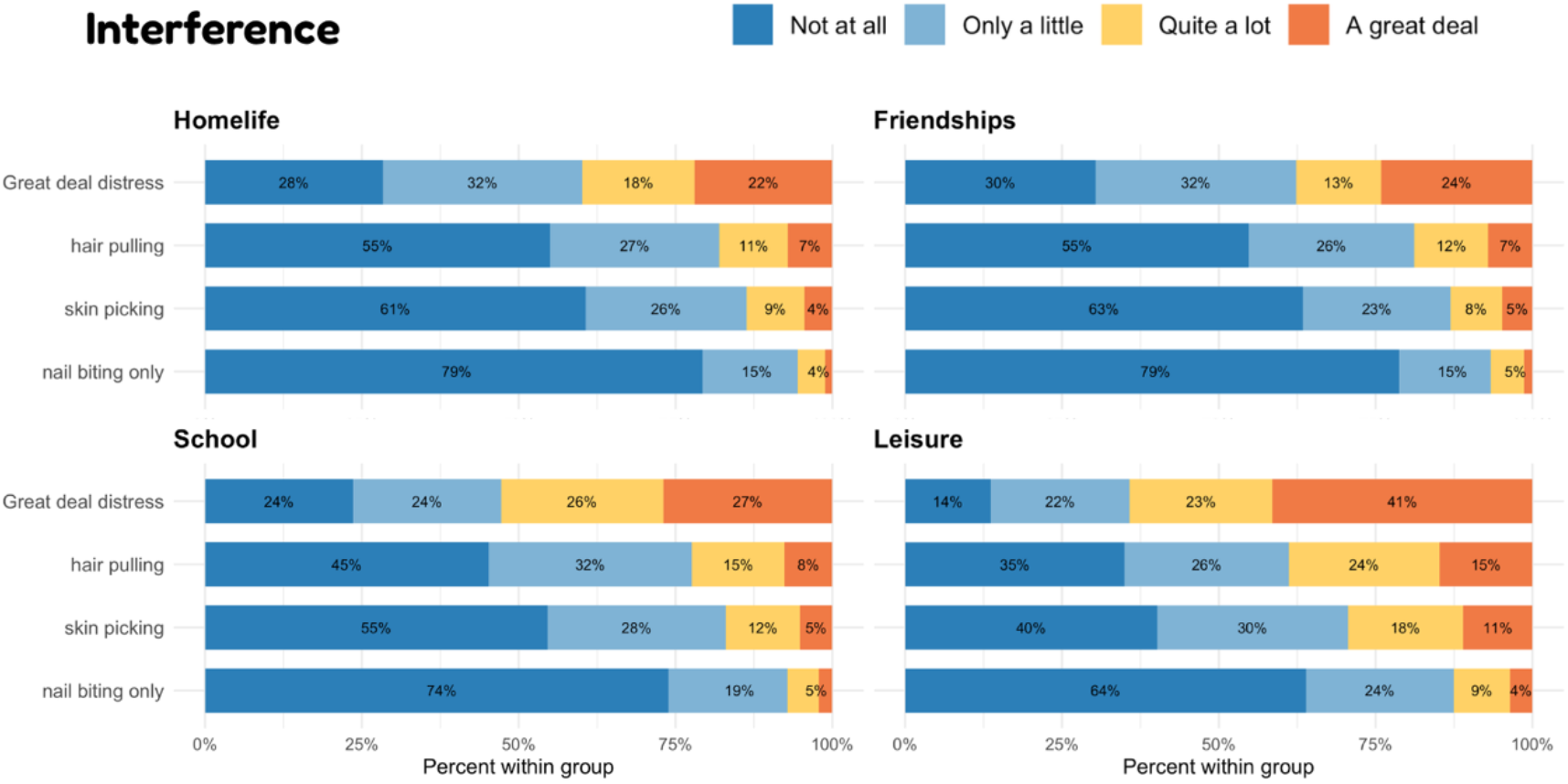
Interference across all four domains of homelife, classroom, friendships and leisure, shown for nail biters only, skin-pickers, hair-pullers and for those with the highest level of distress. Interference can be seen across all domains, particularly in those individuals with greatest distress scores, with the highest proportions being observed in leisure activities.

### Gender differences

There were marked differences in the proportions reporting BFRBs by gender (Figure 3). Overall, 45.6% of 2112 boys/men reported at least one BFRB, compared with 64.6% of 3132 girls/women and 84.2% of 351 TGD participants.

**Figure 3.**
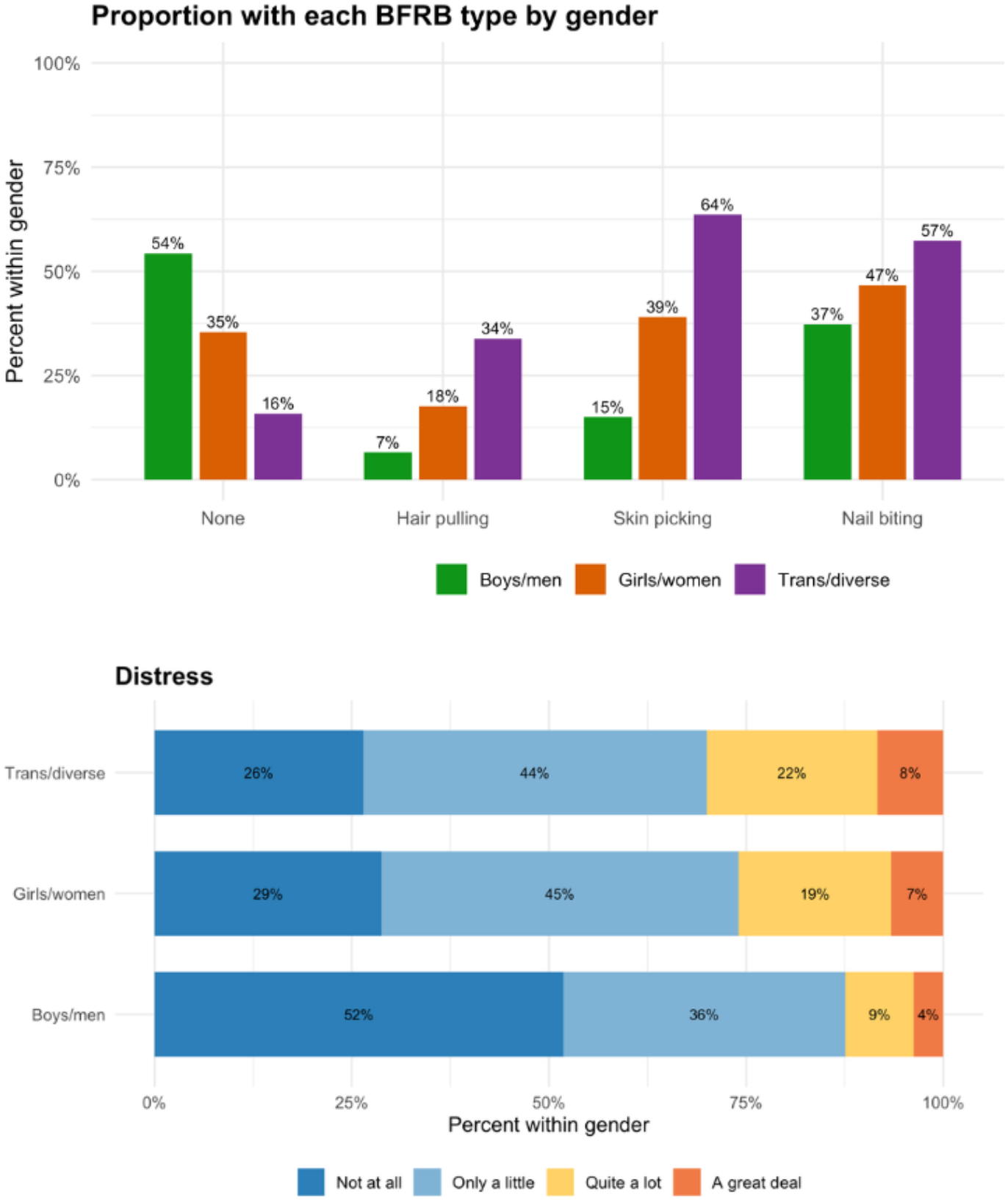
Higher rate of BFRBs and associated distress seen across all BFRBs for girls/women and those identifying as transgender, gender diverse or unsure relative to boys/men.

Across specific behaviours, girls/women and TGD participants consistently reported higher proportions than boys/men. This pattern was particularly pronounced for hair pulling and skin picking. For example, skin picking was reported by 15.0% of boys/men, 39.0% of girls/women, and 63.6% of TGD participants. Nail biting was the most common behaviour across all groups but was also more frequently reported among girls/women (46.6%) and TGD participants (57.4%) than boys/men (37.3%).

Distress associated with BFRBs also varied by gender (Figure 3). Boys/men were more likely to report no associated distress (51.8%) compared with girls/women (28.8%) and TGD participants (26.4%). In contrast, moderate to high levels of distress (“quite a lot” or “a great deal”) were reported by 12.4% of boys/men, 26.0% of girls/women, and 30.0% of TGD participants. The highest level of distress (“a great deal”) was reported by 3.7% of boys/men, 6.6% of girls/women, and 8.4% of TGD participants.

### Associations between BFRBs and self-reported measures of neurodiversity, bullying, and mental health

The proportion of participants reporting problematic BFRBs was higher among those identifying as neurodivergent (Figure 4). Among participants reporting no neurodivergent condition, 10.2% met criteria for problematic BFRBs, compared with 18.4% of those reporting autism, 20.3% of those reporting ADHD, and 20.7% of those reporting both autism and ADHD.

**Figure 4.**
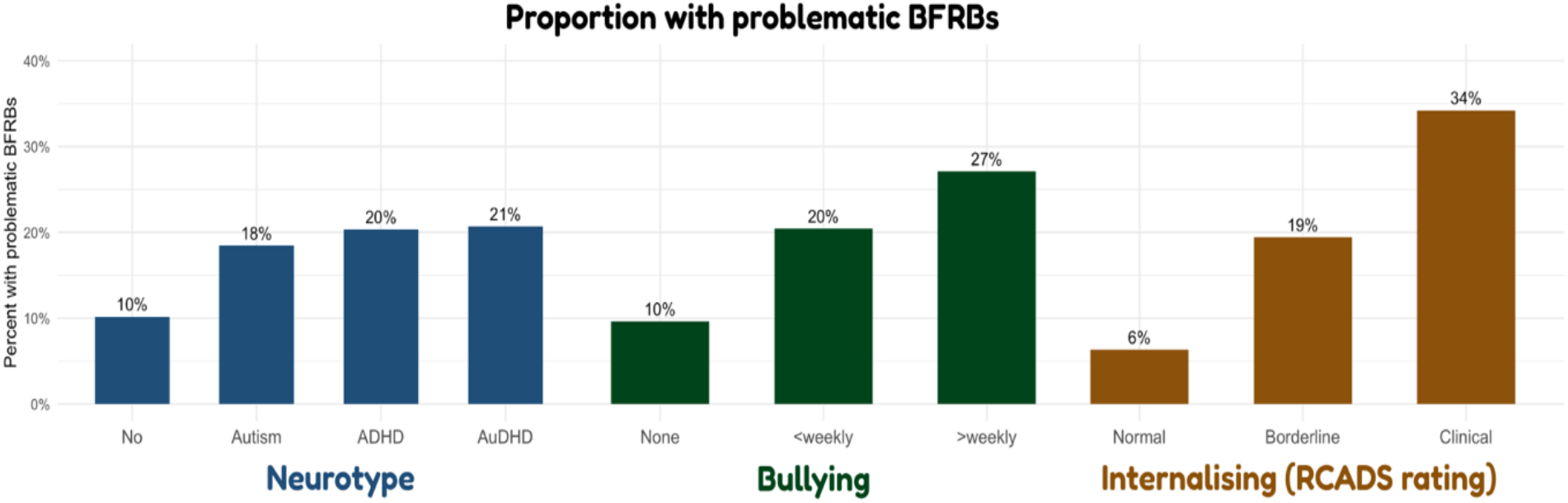
Higher proportion of problematic BFRBs in young people identifying as neurodivergent, experiencing bullying, and/or with higher internalising symptoms (RCADS rating)

Problematic BFRBs were also more common among participants reporting recent bullying (Figure 4). Among those reporting no bullying, 9.6% met criteria for problematic BFRBs, compared with 20.4% of those reporting less frequent bullying and 27.1% of those reporting bullying more than weekly, indicating a dose–response relationship.

Problematic BFRBs were strongly associated with internalising symptoms (Figure 4). Among participants in the normal range of RCADS scores, 6.3% met criteria for problematic BFRBs, compared with 19.4% in the borderline range and 34.2% in the clinical range, indicating a strong gradient across levels of internalising symptoms.

Consistent with this, higher levels of BFRB-related distress were moderately correlated with depressive symptoms (Spearman’s ρ = 0.41, p < .001).

In the multivariable logistic regression model, internalising symptoms emerged as the strongest predictor of problematic BFRBs. Higher RCADS total scores were associated with increased odds of problematic BFRBs (OR = 1.07 per one-point increase, 95% CI 1.07–1.08, p < .001), corresponding to 1.97 per 10-point increase. Gender differences remained after adjustment. Compared with boys/men, girls/women had higher odds of problematic BFRBs (OR = 1.79, 95% CI 1.40–2.29, p < .001), as did participants identifying as TGD (OR = 1.56, 95% CI 1.05–2.32, p = .027). Recent bullying was associated with increased odds of problematic BFRBs, with those reporting bullying more than weekly showing higher odds compared with those reporting no bullying (OR = 1.39, 95% CI 1.01–1.93, p = .046). The association for less frequent bullying was in the same direction but did not reach statistical significance (p = .070). In contrast, neurodivergence was not independently associated with problematic BFRBs after adjustment for other variables. Age was also not significantly associated with the outcome. Overall, the model substantially improved fit compared with the null model, indicating that these variables jointly explained variation in problematic BFRBs.

As a secondary analysis, BFRB severity (none, low/no distress, high distress) was examined using ordinal regression. The pattern of results was consistent with the primary model, with higher internalising symptoms associated with greater severity, and gender and bullying also showing associations with increasing severity. Neurodivergence was associated with severity in this model, suggesting effects across the full spectrum of BFRB severity.

In post-hoc analyses including both depression and anxiety scores, both were independently associated with problematic BFRBs. However, these estimates should be interpreted cautiously given the high correlation between the two measures. Higher autistic trait scores (CATI) were associated with increased odds of problematic BFRBs. In models including both CATI scores and categorical neurodivergence, CATI remained associated whereas neurodivergence was no longer significant, suggesting that dimensional autistic traits accounted for the observed association with neurodivergence.

## Discussion

In this large community survey of school-aged adolescents (n=5,437), 58.5% of students reported experiencing at least one BFRB, with the most common behaviour being nail-biting, followed by skin-picking then hair-pulling. A substantial minority (13% overall, or 22% of those with at least one BFRB) report symptoms as meaningfully distressing. Co-occurrence is the norm, with 86% of hair-pullers and 75% of skin-pickers reporting at least one other BFRB, suggesting that BFRBs should be conceptualised as a multi-behaviour syndrome rather than discrete conditions. Girls and TGD individuals were significantly more likely to both engage in, and be distressed by, BFRBs than boys. The incidence of problematic BFRBs was also significantly associated with internalising symptoms (depression and anxiety) and experiencing recent bullying. Overall, the results confirm that distressing BFRBs are common in adolescents, and neglect of these conditions might be particularly problematic for those who are also struggling with mental health and social challenges.

A notable finding was the substantial co-occurrence of BFRBs within individuals. Nearly half of those reporting any BFRB endorsed more than one behaviour, indicating that these behaviours are not typically experienced in isolation. Co-occurrence also varied by BFRB type: participants reporting hair pulling or skin picking were more likely to report multiple behaviours than those reporting nail biting. Together, these findings suggest that BFRBs may be better conceptualised as manifestations of a shared underlying process that varies in severity, rather than as distinct conditions. This challenges the common tendency in the literature to examine individual BFRBs separately, and may help explain their overlapping associations with internalising symptoms and social stressors [28]. Anecdotally, there is a suggestion of a difference in referral pathways, with skin-picking being more common in dermatology clinics, and hair-pulling more common in psychological services. Developing a shared understanding and language around BFRBs has important implications for how they are screened for, assessed and addressed in clinical and research settings [29].

The proportion of young people reporting distressing BFRBs in our sample was broadly consistent with prior population-based studies using self-report measures, which tend to yield higher rates than clinical or diagnostic studies [3]. Although many participants reported little or no distress associated with their BFRBs, 21% experienced at least moderate distress, and a smaller group (6% of those with BFRBs, 3% of the whole sample) reported a great deal of distress. We found that BFRBs were associated with functional interference across multiple domains, including home life, friendships, leisure, and classroom learning. Interference was greatest among those reporting the highest levels of distress, indicating that distress is a key clinical marker.

Gender differences were evident across both prevalence and distress. Girls/women and TGD participants were more likely to report BFRBs and to experience greater associated distress. Importantly, these differences persisted after adjustment for internalising symptoms and other variables, suggesting that gender differences in BFRBs are not fully explained by differences in anxiety or depression. Our overall proportions reporting at least one BFRB in girls (65% overall) and boys (46% overall) lie in contrast to the previous reports of approximately 4:1 female:male in adult clinical samples [7], [8]. If we use ‘a great deal’ of distress as a proxy for likely clinical need, then the proportion of boys (3.7%) to girls (6.6%) should be around 1:1.8. We also report, for the first time, substantially higher proportions of BFRBs in TGD individuals (84% overall, 8.4% with a great deal of distress), which extends a limited existing literature on BFRBs in gender/sexual minorities [30]. This underscores the need for more inclusive approaches to understanding and supporting people with BFRBs.

The strongest predictor of problematic BFRBs in our analysis was internalising symptoms (RCADS total). We observed a steep gradient across RCADS categories, alongside a moderate correlation between distress and depressive symptoms, which support the view that BFRBs are closely linked to broader emotional difficulties, as reported in previous adolescent and adult samples [10], [17]. This strong association provides support for models that suggest that these behaviours may function as self-regulatory responses to emotional distress [31] [32]. Unfortunately, BFRBs often become chronic sources of suffering in their own right, characterised by loss of control, shame, and concealment [33] [34], creating a vicious cycle of urges and consequences [35], [36].

Bullying was also associated with problematic BFRBs, with a clear dose–response relationship observed in relation to the reported frequency of bullying. Notably, the association persisted after adjustment for internalising symptoms, suggesting that bullying may confer risk through pathways not fully captured by symptom measures alone. This aligns with models in which interpersonal stressors contribute to the emergence or maintenance of repetitive behaviours, potentially through increased self-consciousness, shame, or emotional dysregulation [18], [19] [35]. Interestingly, evidence from non-human primates suggests that self-grooming behaviours are more common in individuals who are experiencing greater ‘social uncertainty’ by virtue of their dominance rank [37]. It may be that self-grooming has a particular soothing function in relation to the discomfort of social isolation and/or ambiguity.

Problematic BFRBs were also more common in neurodivergent young people, although this association was reduced to non-significant after adjusting for internalising symptoms, gender and bullying. This suggests that a higher incidence of BFRBs in neurodivergent young people may be operating through internalising symptoms and/or bullying, which are both common experiences in this population [38], [39]. Ordinal analysis of severity, however, remained significantly associated with neurodivergence, and there was also a significant association with autistic traits. This suggests that dimensional characteristics may better capture the aspects of neurodevelopmental difference relevant to BFRBs than diagnostic categories alone [40].

The major strengths of this study lie in the large sample size of young people who gave ratings of their BFRBs alongside many other measures of mental health and wellbeing. OxWell is conducted through collaborations with schools, and the surveys are carried out in normal school time, increasing the representativeness of the sample relative to typical research cohorts. Nevertheless, there are some aspects of the study that require methodological consideration. The measures included are all self-report, and were not diagnostic interviews, so we do not have information about the clinical status of any of our participants. The sample included in the analyses presented here are those who volunteered to undertake an optional add-on module to the main OxWell survey, so are therefore not fully representative of the full group. As described, this sample included a greater proportion of girls/women and TGD individuals relative to boys, and were slightly younger. They may also have been more motivated and/or able to answer questions more quickly than those who only completed the main survey, which may have influenced reporting and bias the experience of BFRBs reported. Finally, the cross-sectional design precludes any inference about causality. This is particularly important when interpreting the strong association between internalising symptoms and problematic BFRBs, which prior work has indicated is likely to be bi-directional [33], [34] and therefore difficult to disentangle.

To conclude, BFRBs are common in adolescents and co-occurrence is the rule rather than the exception. These findings reveal a substantial and largely unrecognized unmet need; BFRBs are a significant source of distress and impairment, yet are rarely assessed in clinical, school or community settings [41], [42], meaning that young people experiencing clinically meaningful difficulties continue to go unsupported. As such, developing a shared understanding and language around BFRBs and using simple screening questions focused on distress, rather than behaviour alone, could help identify young people who are struggling and open pathways to support [29]. For example: ‘Do you ever pick, pull or bite at your hair, skin or nails in a way that upsets you or gets in the way of things you want to do?’. Interventions that target internalising symptoms and interpersonal sources of stress, such as bullying, are likely to be particularly important for reducing the burden of problematic BFRBs, especially among girls and gender-diverse young people. These behaviours are, by their nature, persistent, and so too must be our efforts to recognise, understand and support those who struggle with them.

## Data Availability

The data used in this study are not publicly available. The study protocol, variable guides, the full list of questions and other supporting information are available on the OxWell Open Science Framework website (https://osf.io/sekhr/). Fully deidentified extracts of the data can be provided to academic research collaborators upon reasonable request after a review process by the research team to ensure that uses of the data fall under the remit of the intended purposes set out in the privacy information and to prevent duplication of analyses.

## Acknowledgements

We gratefully acknowledge the OxWell Study team (https://osf.io/sekhr/), the many students, staff and colleagues in City Councils and service commissioners who contributed their time and experiences. CEM and PW are supported by the NIHR Oxford Health Biomedical Research Centre. The OxWell Student Survey was funded by the NIHR Applied Research Collaboration Oxford and Thames Valley at Oxford Health NHS Foundation Trust, with support from Liverpool City Council and Oxfordshire City Council. The views expressed in this publication are those of the authors and not necessarily those of the National Institute for Health and Care Research or the Department of Health and Social Care.

